# First identification of SARS-CoV-2 Lambda (C.37) variant in Southern Brazil

**DOI:** 10.1101/2021.06.21.21259241

**Authors:** Priscila Lamb Wink, Fabiana Caroline Zempulski Volpato, Francielle Liz Monteiro, Julia Biz Willig, Alexandre Prehn Zavascki, Afonso Luís Barth, Andreza Francisco Martins

**Author notes:** Correspondence Author: Priscila Lamb Wink, LABRESIS- Laboratório de Pesquisa em Resistência Bacteriana, Hospital de Clínicas de Porto Alegre, Rua Ramiro Barcelos 2350, Porto Alegre, RS, Brazil, 90.035-903.

## Abstract

In June 15, 2021, the lineage Lambda (C.37) of SARS-CoV-2 was considered a variant of interest (VOI) by the World Health Organization. This lineage has high prevalence in some South America countries but it was described only occasionally in Brazil. Here we describe the first report of the SARS-CoV-2 Lambda variant in Southern Brazil. The sequence described in this paper presented all the eight C.37 defining lineage mutations (ORF1a gene: Δ3675-3677; Spike gene: Δ246-252, G75V, T76I, L452Q, F490S, D614G, and T859N) in addition to other 19 mutations. Considering that this VOI has been associated with high rates of transmissibility, the possible spread in the Southern Brazilian community is a matter of concern.

---

Since the emergence of the severe acute respiratory syndrome coronavirus 2 (SARS-CoV-2) in December 2019, new lineages of this virus have been described. The recent emergence of SARS-CoV-2 variants of interest (VOI) and variants of concern (VOC) with potentially increased transmissibility and reduced sensitivity to antibody neutralization, may impact the efficacy of strategies to control the COVID-19 pandemic.^1^ A novel VOI within B.1.1.1 lineage, termed C.37 assigned by the World Health Organization (WHO) as “Lambda” in June 15, 2021, was detected in Peru in August 2020 and has been found in 26 countries in America, Europe, and Oceania, drawing international attention for its rapid expansion.^2-5^ Despite the global spreading of the Lambda variant, in Brazil this lineage was reported only in São Paulo state in February 2021.^5^ Here we describe the first report of the SARS-CoV-2 Lambda variant in Southern Brazil.

A young male who has been in Argentina initiated respiratory symptoms compatible with virus infection while returning to his town, which borders Argentina, in Rio Grande do Sul, the southernmost Brazilian state. Four days after the beginning of the symptoms, he tested positive for SARS-CoV-2 by a point of care test. On the following day, he was admitted to a local hospital but due to the worsening of symptoms he was transferred to the intensive care unit of Hospital de Clínicas de Porto Alegre, a tertiary-care, COVID-19 reference hospital in the state, two days after hospital admission. Oro/nasopharyngeal swabs were collected and real-time reverse transcription-PCR testing for two genes of the nucleocapsid protein (N1 and N2) for SARS-CoV-2 was carried out,^6^ confirming the SARS-CoV-2 infection. The specimen (203_LABRESIS) was submitted to the whole-genome sequencing (WGS) as part of an epidemiological study. Sequencing libraries were prepared using the CleanPlex SARS-CoV-2 panel protocol (Paragon Genomics, Hayward, United States) for target enrichment and library preparation, following manufacturer instructions (https://www.paragongenomics.com/wp-content/uploads/2020/03/UG4001-01_-CleanPlex-SARS-CoV-2-Panel-User-Guide.pdf). The resulting libraries were sequenced in Illumina MiSeq (Illumina, San Diego, US). Consensus sequences were generated by the QIASeq SARS-CoV-2 pipeline (QIAGEN CLC Genomics Workbench 21, Germantown, United States) with high quality whole-genome sequence (average coverage 3,142, <2% Ns, 29,873 Kb). The specimen 203_LABRESIS was classified as C.37 variant using the Phylogenetic Assignment of Named Global Outbreak Lineages (Pangolin) software tool (v3.0.2)^3^ and the sequence was deposited into the GISAID database^5^ under the number EPI_ISL_2617911.

The Lambda variant (C.37) is defined by a deletion in the ORF1a gene (Δ3675-3677), also present in Alpha (B.1.1.7, United Kingdom), Beta (B.1.351, South African), and Gamma (P.1, Brazil) VOCs. In addition, the Lambda variant displays a novel deletion and multiple nonsynonymous mutations in the spike gene (Δ246-252, G75V, T76I, L452Q, F490S, D614G, and T859N).^2,4^ The mutations L452Q and F490S are mapped in the receptor-binding domain (RBD) region and the F490S has been associated with reduced susceptibility to antibody neutralization.^1^ The isolate 203_LABRESIS presented all the eight C.37 defining lineage mutations described above in addition to other 19 mutations that have already been described in members of this lineage (Fig. 1).^2,4,7^ The 203_LABRESIS sequence was aligned with high quality and coverage complete genome sequences from Brazil (1), Argentina (16), Chile (24), and Peru (26) available in EpiCoV database in GISAID to perform a comparative phylogenetic tree. The 203_LABRESIS sequence clustered together with another C.37 variant sequence from Argentina identified in February, 2021 (Fig. 1).

**Figure 1.**
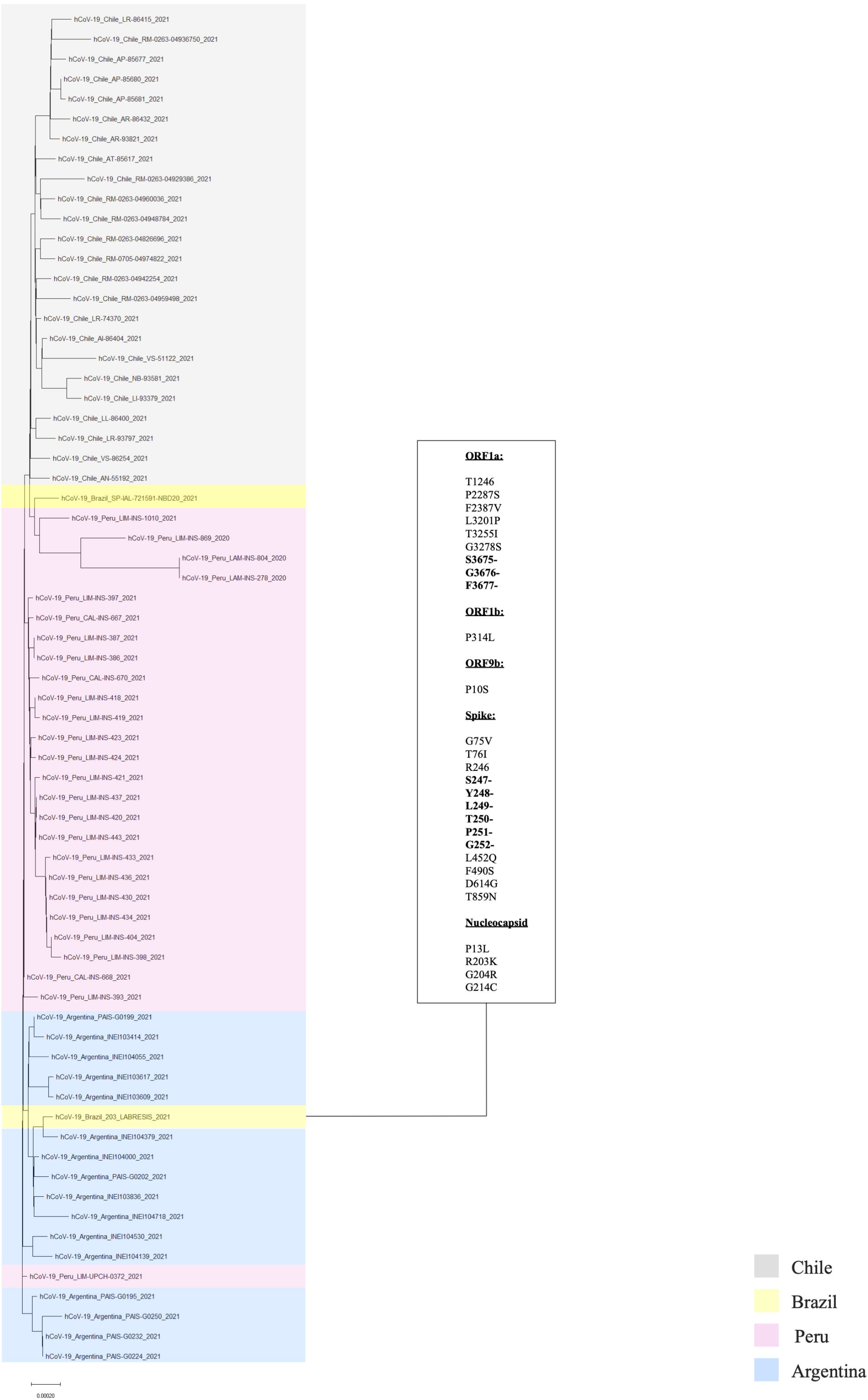
Maximum likelihood phylogenetic tree (ML, GTR, 1,000 bootstrap) of the C.37 lineage. The phylogenetic tree was constructed using the 203_LABRESIS sequence obtained from this study, 16 sequences from Argentina, 24 from Chile, 26 from Peru and 1 from São Paulo State of Brazil. All sequences were available in EpiCoV database in GISAID. This dataset was aligned using MAFFT v7.475 and subjected to maximum likelihood (ML) phylogenetic analysis using IQ-TREE v2.1.2 (GTR+F+R4, SHaLRT with 1,000 replicates). Branches are colored according to the countries (Chile in grey, Brazil in yellow, Peru in pink, and Argentina in blue). The scale of the phylogenetic branches is given as substitutions per nucleotide site. All mutations identified in the 203_LABRESIS sequence (deletions in the ORF1a and Spike genes are shown in bold) are listed in the box.

The high prevalence of the Lambda variant has been described particularly in South America countries such as Chile, Peru, Ecuador, and Argentina, where this new VOI is spreading rapidly, being associated with substantial rates of community transmission.^2^ It is believed that the health care system critical situation and the recent report of increased deaths in these countries is associated with the rising prevalence of the Lambda variant.^8-10^

Noteworthy, only in June 15, 2021, Lambda was considered a VOI by the World Health Organization.^2^ It is not known whether the Lambda variant is more transmissible or more pathogenic than other variants or whether it is able to evade the effect of available vaccines. The novel S: Δ246-252 deletion and additional mutations in the spike protein should be taken into account to understand their effects on viral fitness and host interaction. This first report of SARS-CoV-2 Lambda variant in Southern Brazil raises concern regarding a possible dissemination of this lineage in the region. Moreover, considering that this VOI has rapidly spread in Peru, Ecuador, Chile, and Argentina, we believe that Lambda variant has a considerable potential to become VOC.

## Data Availability

The datasets generated during and/or analyzed during the current study are available from the corresponding author on reasonable request.

## Financial support

This study was funded by FAPERGS (20/2551-0000265-9), National Institute of Antimicrobial Resistance Research - INPRA (MCTI/CNPq/CAPES/FAPs nº 16/2014) and by Fundo de Incentivo a Pesquisa e Eventos do Hospital de Clínicas de Porto Alegre (FIPE/HCPA) (Project no. 2020-0163). PLW and FCZV were supported by a grant from the “Coordenação de Aperfeiçoamento de Pessoal de Nível Superior (CAPES)”.

## Conflict of interest

APZ, ALB and AFM are research fellows of the National Council for Scientific and Technological Development (CNPq), Ministry of Science and Technology, Brazil. APZ has received a research grant from Pfizer not related to this work. All other authors have no conflict to declare.

## Acknowledgements

We thank the staff of “Laboratório de Diagnóstico de SARS-CoV-2” as well as the staff of the Hospital Infection Committee of our institution (“Hospital de Clínicas de Porto Alegre”) for providing data used in this study. This research has been conducted using the “Hospital de Clínicas de Porto Alegre” Biobank resources under Application Number GPPG 2020-0163.

## Ethical statement

This study was approved by the Ethics Committees from Hospital de Clínicas de Porto Alegre (CAAE: 30767420.2.0000.5327).

## Notes

### Competing Interest Statement

The authors have declared no competing interest.

### Clinical Trial

Not Applicable

### Author Declarations

This study was approved by the Ethics Committees from Hospital de Clinicas de Porto Alegre (CAAE: 30767420.2.0000.5327).

